# An Integrated Data-Driven Model for Clinical Phenotyping of Tuberculosis Disease Severity

**DOI:** 10.1101/2025.11.25.25341018

**Authors:** Samantha Malatesta, Karen R Jacobson, C Robert Horsburgh, Maha Farhat, Tara Carney, Krista J Gile, Eric D Kolaczyk, Laura F White

## Abstract

A common approach to describing tuberculosis (TB) disease severity is to use a binary classification such as “advanced” and “minimal or early disease,” though this may not fully capture the range of clinical presentations. As individuals transition through stages of disease, we expect to observe increased bacterial burden and inflammation which corresponds to worsening disease severity and increased risk of a negative outcome. We develop a new method, tuberculosis SeveriTy Assessment Tool for Informed Stratification (TB-STATIS), to understand the various disease severity phenotypes that exist at time of clinical presentation. Our method integrates data from multiple sources (i.e. smear microscopy, chest x-ray findings, symptoms, etc.) to identify a set of disease severity classes and obtain a predicted disease class for each individual given their observed data. Our approach is motivated by the statistical framework used in event-based modeling, a type of data-driven disease progression modeling. We show in simulation TB-STATIS can correctly identify the true set of disease classes with various sample sizes, data sources to integrate, and levels of uncertainty in the observed data. We apply TB-STATIS to two data sets, data from an observational TB cohort in South Africa and data from a global phase 3 clinical trial that tested the non-inferiority of two 4-month regimens compared to the standard 6-month regimen for the treatment of TB. We observe disease classes generated from TB-STATIS correlate with culture conversion, a proxy for TB treatment response. We demonstrate our approach to classifying TB disease severity generates clinically meaningful strata.

## 1 Introduction

The current standard treatment for TB is effective but requires multiple antibiotics taken daily for 6 months. Cure rates remain sub-optimal globally due to challenges with late diagnoses, treatment adherence, and drug side-effects as well as lack of sterilization [1]. Efforts to improve outcomes and shorten therapy continue [2]. A recent trial showed that a 4-month regimen for drug-susceptible TB was non-inferior to standard treatment [3]. Secondary analyses using simple risk-stratification based on Xpert MTB/RIF cycle threshold and chest radiography linked disease phenotypes to unfavorable outcomes [4]. Similarly, pooled patient-level analyses across several trials found that individuals with non-cavitary disease or low sputum burden could achieve relapse-free cure with 4-month therapy [5]. These findings highlight the growing potential for stratified treatment based on disease severity at diagnosis.

When evaluating individuals for TB disease, we routinely collect diagnostics that reflect disease severity, including smear grade, time to culture positivity, nucleic acid amplification tests (NAATs), chest-radiography findings, and symptom severity. Active TB lies on a spectrum from minimal disease (asymptomatic, minimal radiographic changes, bacteriologically negative) to extensive disease (symptomatic, smear-positive, cavitary radiographic abnormalities) [6]. Progression along this spectrum generally corresponds to worsening severity and higher risk of adverse outcomes. Data-driven disease progression modeling (DDPM) provides methods to reconstruct disease timelines directly from observed data [7], and the event-based model (EBM), a major DDPM framework, has been widely applied to neurodegenerative diseases [8–10]. Motivated by this framework, we aim to develop a model that integrates available TB diagnostic data to describe an ordered spectrum of disease severity.

The EBM provides a probabilistic framework for individual disease staging by estimating the population-level sequence in which biomarkers transition from normal to abnormal. It assumes that biomarkers become increasingly abnormal as disease progresses, allowing the event sequence to be inferred from cross-sectional data alone [7]. This is well suited to TB for two reasons: (1) we typically only observe individuals at diagnosis before treatment begins, and (2) the model does not require clinical outcomes to estimate the disease sequence. We are often interested in multiple TB outcomes (i.e. treatment success, time to sterilization, and relapse) and being able to use a single model without retraining for each outcome is a major advantage.

We introduce TB-STATIS (Tuberculosis SeveriTy Assessment Tool for Informed Stratification), a data-driven approach that integrates multiple diagnostic measures such as sputum burden, symptoms, and chest X-ray, to classify TB disease severity at diagnosis. While disease severity cannot be measured directly, we assume the collected can inform severity classes. We propose a model that integrates the statistical frameworks from two extensions of the EBM, resulting in an approach tailored to the types of data typically encountered in TB research [11,12]. It requires only cross-sectional data and is flexible, accommodating different diagnostic algorithms and variable data granularity across studies and programs. The model likelihood is adapted for categorical and ordinal inputs, reflecting the nature of TB diagnostics, and it does not require control data due to the use of validated diagnostic measures with pre-defined cutoffs. To demonstrate clinical utility, we correlate disease severity classes with mycobacterial culture sterilization and unfavorable outcomes using data from a phase 3 clinical trial and an observational TB cohort.

## 2 TB-STATIS for Disease Severity Classification

### 2.1 Data Structure and Notation

Our method requires individual level data collected at time of TB diagnosis that are associated with disease severity (i.e. smear microscopy, culture, symptoms, chest x-ray, etc.). Our input data is the set of clinical measures *X* = {*x*_1_, …, *x*_*I*_} collected on individuals *j* = 1, …, *J*. Each measure *x*_*i*_ has a set of clinical states *R* = 0, …, *W*_*i*_ where *W*_*i*_ + 1 is the number of clinical states for clinical measure *x*_*i*_ and more advanced clinical states correspond to more severe disease. Each individual occupies exactly one clinical state for each clinical measure. We denote clinical state *R* within clinical measure *x*_*i*_ as *x*_*i,R*_. For example, if clinical measure *x*_1_ is self-reported cough, then *x*_1_has two clinical states (R = 0, 1) which are *x*_1,0_ and *x*_1,1_ corresponding to no cough and cough, respectively. Individuals in clinical state *x*_1,1_ that report a cough have more severe disease than those in state *x*_1,0_ that do not report a cough.

Given input data *X* = {*x*_1_, …, *x*_*I*_} where each *x*_*i*_ is a clinical measure, the parameter of interest we want to estimate is the disease sequence *S* which is an ordered list of sets *S* = {*s*(*k*), *k* = 1, …, *l*} describing classes 1, …, *l* of increasing disease severity. Each *s*(*k*) contains one or more clinical states that belong to disease class *k*, and these clinical states originate from the different clinical measures in the input data. We allow clinical states for a single measure *x*_*i*_ to occur simultaneously in the same class *k* in sequence *S* if the clinical states are consecutive in *x*_*i*_. At class 0 which is the set of clinical states that are least severe for each clinical measure (i.e. *R* = 0), *k* = 0. For TB-STATIS, we assume that clinical states are monotonically non-decreasing in *S* and disease severity worsens with more advanced clinical states. We depict an overview of TB-STATIS in Figure 1A.

**Figure 1.**
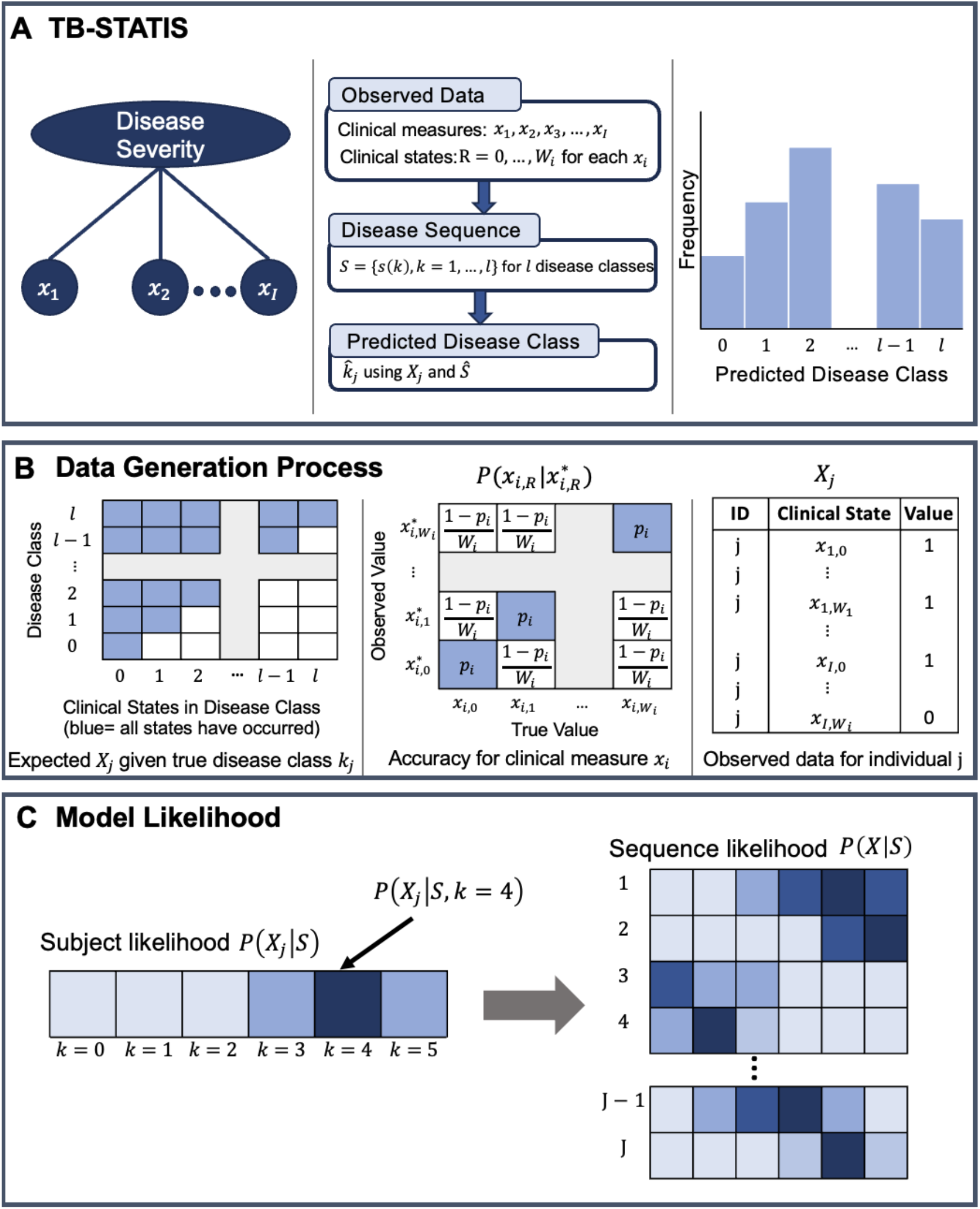
Overview of TB-STATIS, the underlying data generation process, and the model likelihood. Panel A shows an overview of the model from conceptualizing disease severity as a latent variable (left) to estimating a disease sequence and predicting a disease class for each individual (middle and right). Panel B shows the data generation process where the observed data depends on the true disease class for an individual and the accuracy of each clinical measure. Panel C shows an example of the subject specific likelihood given the observed data *X*_*j*_ for the *j*_*th*_ individual (left). The sequence likelihood is calculated from all subject specific likelihoods (right). Each square is a probability with darker shades representing higher probabilities.

An example sequence for three clinical measures: cough, infiltrates, and smear microscopy (Table 1) is *S* = {(*x*_3,1_), (*x*_1,1_, *x*_2,1_), (*x*_1,2_)}. In this sequence, there are three disease classes, cough occurs in class 1, unilateral infiltrates and smear positivity occur in class 2, and bilateral infiltrates occur in class 3. Class 0 contains the clinical states (*x*1,0, *x*2,0, *x*3,0).

**Table 1.** Example of clinical states and their corresponding disease class.

| Clinical Measure | Clinical State | Disease Class |
| --- | --- | --- |
| Infiltrates ($x_1$) | Bilateral ($x_{1,2}$) | 3 |
| | Unilateral ($x_{1,1}$) | 2 |
| | None ($x_{1,0}$) | 0 |
| Smear microscopy ($x_2$) | Positive ($x_{2,1}$) | 2 |
| | Negative ($x_{2,0}$) | 0 |
| Cough ($x_3$) | Yes ($x_{3,1}$) | 1 |
| | No ($x_{3,0}$) | 0 |

### 2.2 Model Specification

We assume two underlying processes that generate the observed data which we depict in Figure 1B. First, if an individual is in class *k* in the model, then clinical states in classes 1, …, *k* have occurred and clinical states in classes *k* + 1, …, *l* have not occurred (Figure 1B, left). Second, at the individual level, there may be differences between the data we observe and the data we expect given an individual’s disease severity class. This variability can manifest for numerous reasons. If the data we are using to estimate the model are self-reported such as symptom screen data, there is the possibility of under or overreporting. Similarly, if we are using laboratory-based diagnostics, false positive or false negative results are a possibility. We formally quantify this variability as the accuracy of each clinical measure 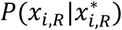 which is the probability the true value is *x*_*i,R*_ given we observed 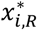 (Figure 1B, middle). For simplicity, we assume 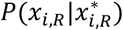 is the same for all clinical states in measure *x*_*i*_ and simplify notation for 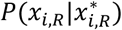 to be 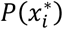.

Using the process described above, we can write the likelihood of an individual’s data (Figure 1C) given disease severity class *k*, sequence *S*, and vector of clinical measure accuracies 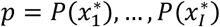, which is user-specified, as the probability that the set of clinical states *U*_1≤*m*≤*k*_ *s*(*m*) have occurred and *U*_*k*+1≤*m*≤*l*_ *s*(*m*) have not occurred for individual *j*

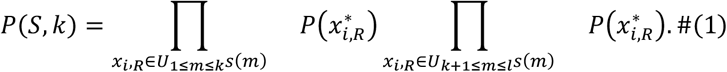

Since the subject specific likelihood depends on *k* which we do not know, we integrate out *k* by taking a weighted sum across all possible classes assuming each disease class is equally likely

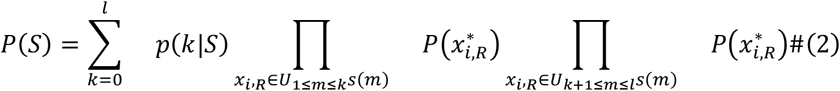

where *l* is the number of disease classes in *S* and 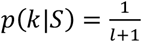.

To obtain the complete likelihood for a given sequence *S*, we assume individuals are independent:

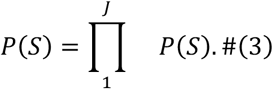

Substituting equation (2) into equation (3), the sequence likelihood is:

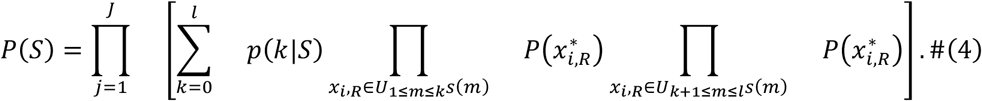

Several key assumptions enable pooling cross-sectional data to estimate a sequence of clinical states. First, we assume TB disease severity sequencing applies to all individuals, allowing inference from a pooled cohort. Second, clinical states within a measure occur sequentially as disease worsens (e.g., an individual reporting a cough must have passed through the “no cough” state). Third, multiple states for a measure can occur simultaneously within the same disease class (e.g., unilateral and bilateral infiltrates). Finally, we assume clinical measures are independent; for instance, smear microscopy results are assumed independent of chest x-ray findings (Table 1).

### 2.3 Model Estimation

We use maximum likelihood estimation to identify the most likely sequence 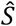 given our observed data. With only a few clinical measures, it is feasible to enumerate all possible sequences and compute 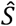 directly. However, including multiple data sources creates too many candidate sequences to enumerate. We adopt an approach similar to Fonteijn et al. and subsequent EBM variations [8,9,11], outlined in Figure 2. We initialize a sequence with clinical states in monotonically non-decreasing order. For a set number of iterations, we propose a nearby candidate sequence and accept it if the log-likelihood increases. With sufficient iterations, this converges to at least a local maximum, and repeating the process with multiple initializations improves convergence toward the global maximum. The key step is proposing candidate sequences, inspired by iterated local search for combinatorial optimization [13]. We perturb the current sequence by swapping two clinical states while maintaining monotonic order, then determine which states occur simultaneously (Figure 2). Further details and an example of sequence perturbation are provided in the Supplementary Material.

**Figure 2.**
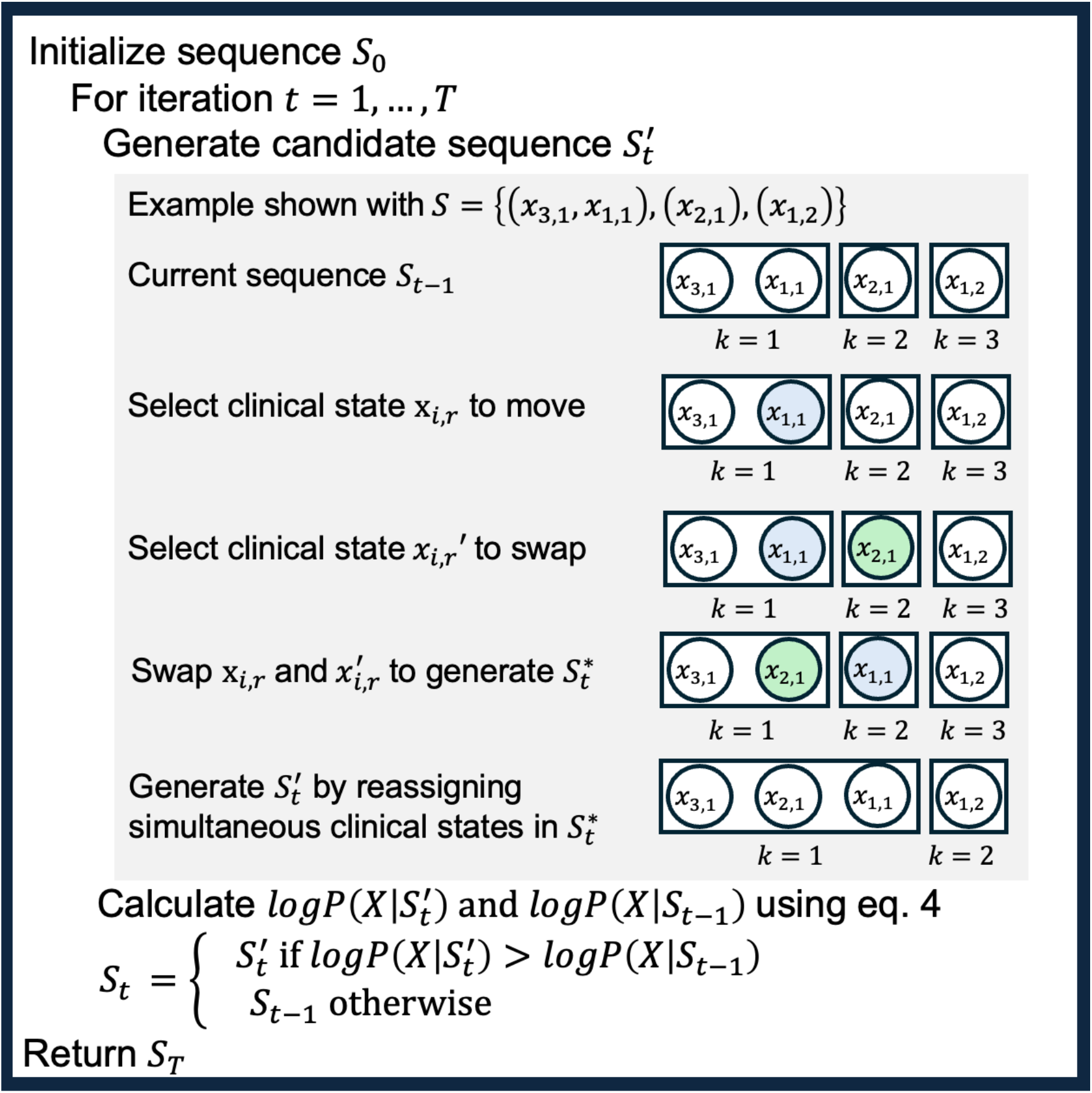
Iterative algorithm for searching for the maximum likelihood sequence. This process is repeated multiple times for different initialized sequences to estimate the maximum likelihood sequence for TB-STATIS.

### 2.4 Model Uncertainty

To quantify uncertainty about 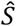 we use bootstrap resampling. We generate a single bootstrap data set by resampling *J* individuals with replacement from our original data. We repeat this process *v* times to generate *v* data sets of size *J*. For each data set we estimate the maximum likelihood sequence. We construct a positional variance diagram (PVD) using our set of estimated sequences 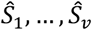 to visualize the uncertainty for each disease class in 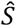 [8]. We expect most of our bootstrap sequences to be identical to 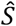. The PVD is a matrix that shows how closely our bootstrap estimated sequences align to 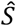. For clinical state *m* and disease class *k*:

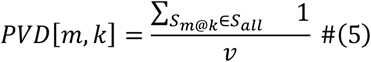

where *S*_*m*@*k*_ denotes a sequence with clinical state *m* at class *k* and *S*_*all*_ is the set of sequences *S*_1_, …, *S*_*v*_ estimated for *v* bootstrap resamples.

### 2.5 Predicted Disease Class

Given an estimated disease sequence, we can calculate the most likely disease class for each individual in our data. For each individual we take the disease class that best fits the data given 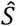:

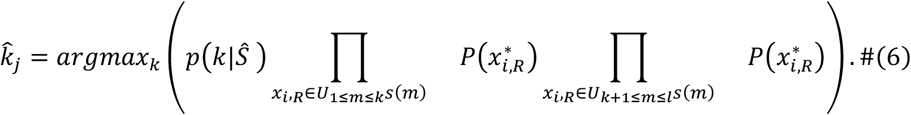

This information allows us to assess the distribution of disease severity at diagnosis, compare severity between individuals to identify associated attributes, and use predicted disease class to explore correlations with clinically relevant outcomes.

### 2.6 Model Fit

To quantify the performance of TB-STATIS for a given data set, we summarize the difference between our observed data and the data we predict given our estimated sequence 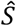. For each clinical state, we observe a vector 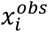 of length *J* where each entry is 1 if clinical state *x*_*i*_ has occurred and 0 otherwise for individual *j*. Similarly, using the predicted disease class 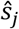, we can calculate the predicted data for clinical state *x*_*i*_ as the vector 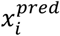 where each entry is the predicted value for *x*_*i*_ for individual *j* given predicted disease class 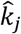 and 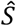. To quantify the similarity between 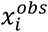 and 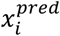, we use the normalized Hamming distance which compares the similarity of two binary vectors by summing the number of positions at which the corresponding elements of two vectors differ [14]:

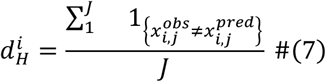

where 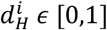 and lower values of 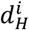 indicate a higher similarity between 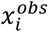 and 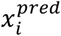. Using equation 7, we obtain 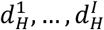 for clinical states *x*_1_, … *x*_*I*_ and calculate the overall similarity for our observed and predicted data:

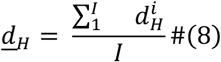

where *<u>d</u>*_*H*_ *ϵ* [0,1] and lower values of *<u>d</u>*_*H*_ indicate a higher similarity and better model fit.

## 3 Simulation Study

### 3.1 Study Design

We conducted a simulation study to evaluate TB-STATIS performance. Data sets varied by size (M = 100, 250, 500) and number of clinical states (I = 4, 8, 12), and we varied the accuracy of each clinical state, 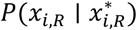, as defined in Section 2.2. For simplicity, we set the same value *p* for all clinical states within a simulated dataset (*p* = 0.75,0.85,0.95) and generated 1,000 datasets for each combination of *M, I*, and *p*. True disease sequences were generated assuming all sequences equally likely, and data were simulated using the process in Section 2.2. TB-STATIS was estimated for each dataset using 10 different initial sequences and 2,000 iterations each, with *p* set to the true value used to generate the data. To assess sensitivity, we re-estimated TB-STATIS using *p* values 0.10 lower than the true values. We also tested the independence assumption by simulating correlated clinical measures with correlations ranging from 0.2 to 0.8.

For all simulations, we quantify differences between the true and estimated disease sequences using the Kendall’s tau distance, which adjusts for the number of clinical states [15]. Values range from 0 to 1, with values near 0 indicating closer agreement between the true and estimated sequences. For each setting, we report the proportion of datasets with a Kendall’s tau distance of 0, indicating exact recovery of the true sequence, and visualize the distribution of distances greater than 0. We also evaluate performance at the disease class level by predicting each individual’s class from the estimated sequence and calculating the Kendall’s rank correlation between true and predicted classes within each dataset.

### 3.3 Results

We present results for estimating sequences of disease severity classes across different simulated settings in Figure 3. Overall, TB-STATIS performs best with fewer clinical states, higher clinical state accuracy (*p*), and larger sample sizes, as indicated by a higher proportion of datasets where the Kendall’s tau distance equals 0. For most settings with 4 or 8 clinical states and *p* ≥ 0.85, TB-STATIS recovers the true disease sequence in 90% or more of simulated datasets. For settings with 12 clinical states, the proportion of datasets with Kendall’s tau distance of 0 is substantially lower, though the distribution of distances greater than 0 has a median of ∼0.1, suggesting that the maximum likelihood sequence is still generally similar to the true sequence. Figure 4 summarizes the Kendall’s tau correlation between true and predicted disease classes within each dataset. Higher values of *p* correspond to stronger positive correlations, indicating good agreement between predicted and true classes at the individual level. When *p* = 0.85 or 0.95, the median correlation exceeds 0.75, regardless of the number of clinical states.

**Figure 3.**
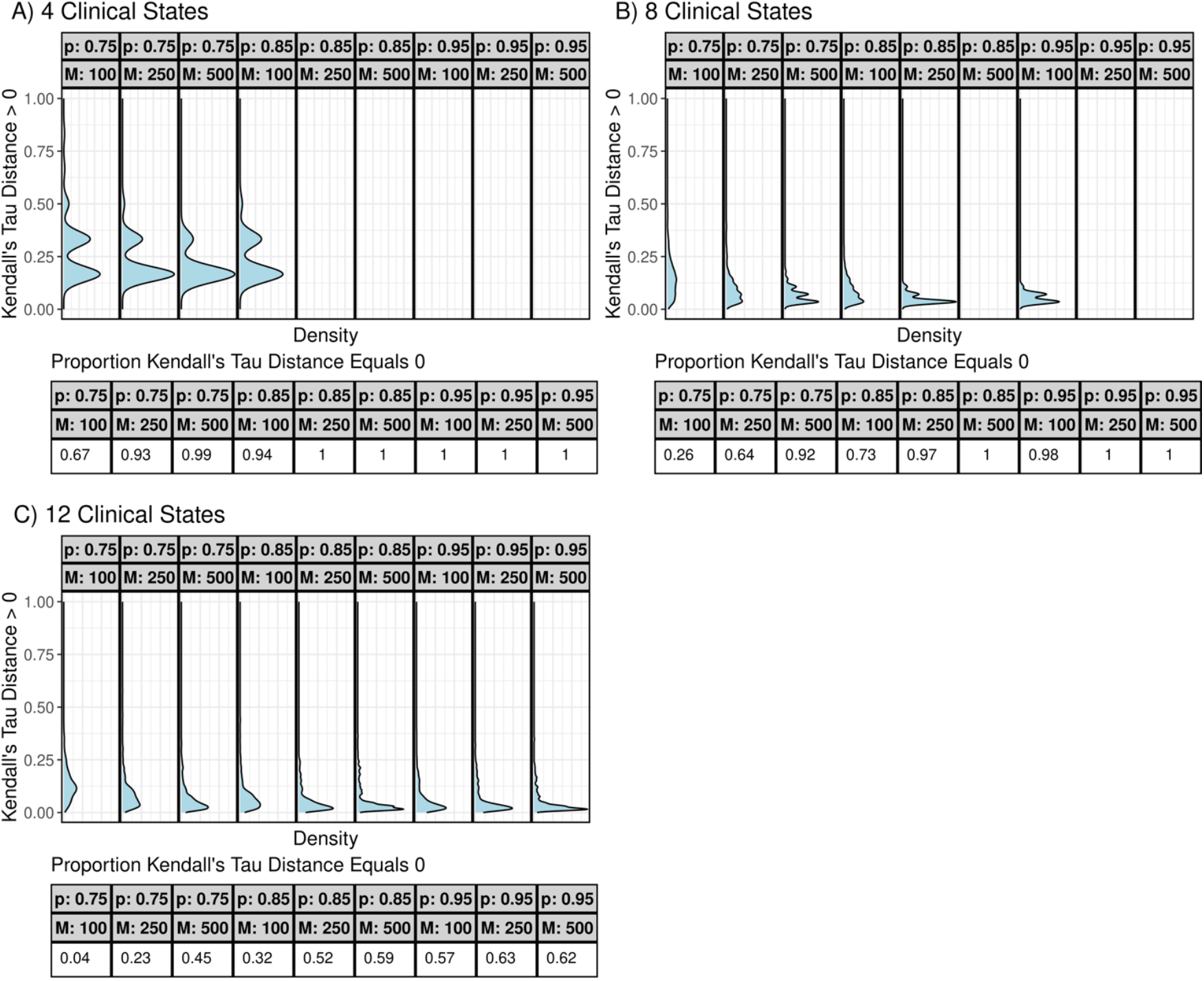
Kendall’s tau distance for estimated and true sequences across simulated settings. For each setting, we summarize the proportion of simulated data sets where the sequence estimated by TB-STATIS is identical to the ground truth sequence (Kendall’s tau distance = 0). These proportions are summarized in the table below each plot in each panel. For simulated data sets where the Kendall’s tau distance is greater than 0, we present the distribution as a density plot. When the proportion of simulated data sets with Kendall’s tau distance equal to 0 is 1, no density plot is presented.

**Figure 4.**
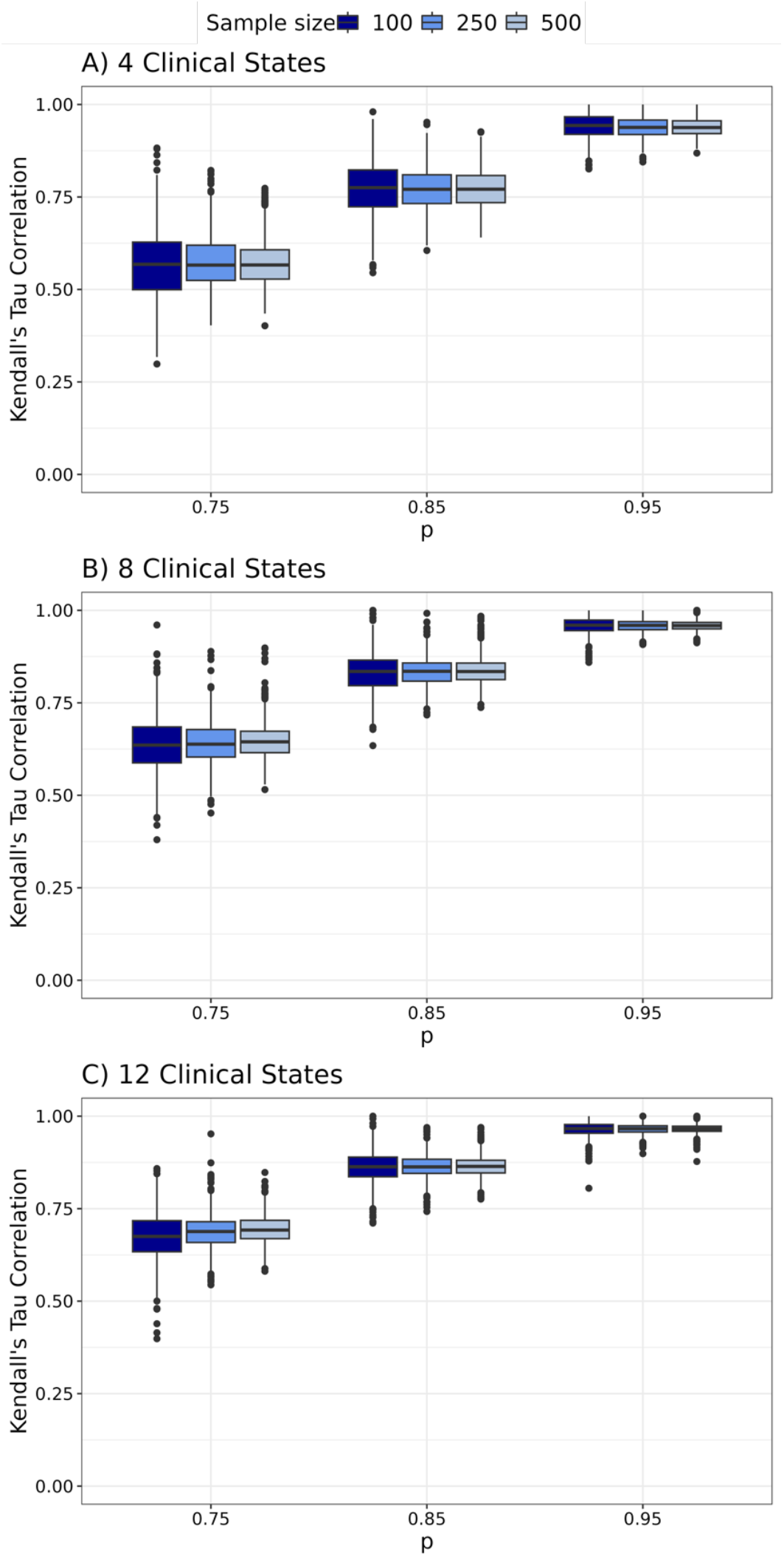
Kendall’s tau correlation quantifying the agreement between predicted and true disease class at the individual level. Box plots represent the distribution of the correlation between true and predicted disease class across simulated settings.

Results from the sensitivity analyses are provided in the Supplementary Material (Figures S1-S5). When the user-specified value for *p* is 0.1 lower than the true value, results are similar to those obtained using the correct *p*. For 4 or 8 clinical states with *p* ≥ 0.85, TB-STATIS correctly estimates the disease sequence in ≥ 90% of simulations (Figure S1). Performance declines for 12 clinical states or *p* = 0.75. At the disease-class level, correlations between true and predicted classes show a similar pattern (Figure S2). When clinical measures are correlated, performance patterns are consistent across sample sizes and *p* values, though overall accuracy decreases with stronger correlation. TB-STATIS remains effective for up to 8 states when *p* ≥ 0.85 (Figures S3–S5).

## 4 Disease Severity and TB Treatment Outcomes

Individuals with more severe TB typically experience longer times to sterilization and higher risk of unfavorable outcomes. TB-STATIS predicts disease severity class from clinical data collected at diagnosis. We demonstrate its ability to stratify individuals into clinically meaningful classes by correlating predicted severity with TB outcomes in two datasets. In Section 4.1, we apply TB-STATIS to data from the Rapid Evaluation of Moxifloxacin in Tuberculosis (REMoxTB) study (NCT00864383), correlating disease class with both unfavorable outcomes and TB sterilization. In Section 4.2, we apply it to data from the Tuberculosis Treatment and Alcohol Use Study (TRUST) (NCT02840877), correlating predicted severity with TB sterilization. Additionally, in the TRUST cohort, we examine the relative frequencies of attributes known to associate with advanced TB across severity classes to further confirm that TB-STATIS generates meaningful strata.

### 4.1 Disease Severity in the REMoxTB Trial

The REMoxTB study was a large, randomized, placebo-controlled Phase III trial designed to test the non-inferiority of two Moxifloxacin-containing 4-month regimens compared to the standard 6-month regimen for TB treatment [16]. REMoxTB data are publicly available from the Platform for Aggregations of Clinical TB Studies (https://c-path.org/tools-platforms/tb-pacts/). We include data on 1,351 participants. Using clinical and laboratory measures collected at randomization (Table 2) and their corresponding clinical states, we applied TB-STATIS to predict a disease severity class for each participant. We then correlated predicted severity with two clinical outcomes: (1) unfavorable TB-related outcome, defined as bacteriologically or clinically confirmed failure or relapse, and (2) 8-week culture status. Further details and additional results are provided in the Supplementary Material.

**Table 2.** Clinical measures in the REMoxTB trial and the estimated disease sequence from TB-STATIS.

| Data source | Clinical Measure | Clinical State | $P(x_i\vert x_i^*)$ | $\hat{S}$ |
| --- | --- | --- | --- | --- |
| Chest x-ray | Cavitary disease | Yes | 0.95 | 1 |
|  |  | No |  | 0 |
| Sputum | Smear microscopy | 4+ | 0.95 | 4 |
|  |  | 3+ |  | 3 |
|  |  | 2+ |  | 2 |
|  |  | 1+ |  | 1 |
|  |  | Negative |  | 0 |
| Symptoms | Cough | Yes | 0.85 | 1 |
|  |  | No |  | 0 |
|  | Fever | Yes | 0.85 | 1 |
|  |  | No |  | 0 |
|  | Night sweats | Yes | 0.85 | 5 |
|  |  | No |  | 0 |
|  | Unexplained weight loss | Yes | 0.85 | 1 |
|  |  | No |  | 0 |

We estimated 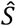 to have six disease classes using our observed data and specified values for 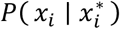 (Table 2). Using 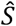, we predicted a disease class for each participant. The distribution is left-skewed, with earlier stages less represented and roughly 50% of participants assigned to the most advanced class (class 5) (Figure 5A). We observe a positive trend in log odds ratios between disease class and 8-week culture positivity, indicating higher odds of positivity with increasing severity (Figure 5B). No clear trend is seen for unfavorable TB-related outcomes, though all disease classes show increased odds compared to the reference group, class 0 (Figure 5B).

**Figure 5.**
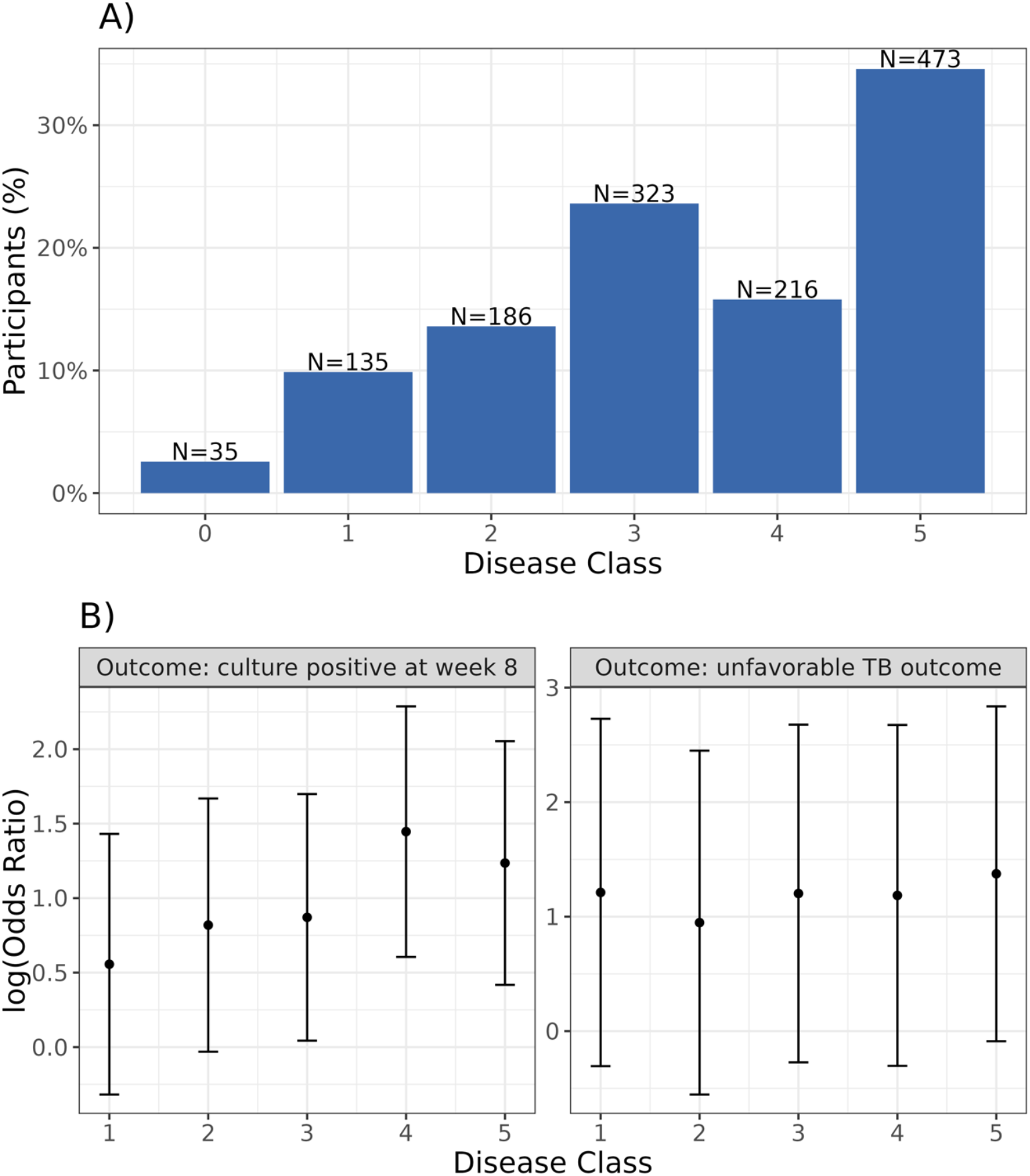
Disease severity classes generated by TB-STATIS and associations with TB treatment outcomes in the REMox cohort. Panel A shows the distribution of predicted disease severity classes. Panel B (left) shows the log odds ratios for the logistic regression model with 8-week culture positivity as the outcome and disease severity as a categorical predictor. Panel B (right) shows the log odds ratios for the logistic regression model with unfavorable TB outcome as the outcome and disease severity as a categorical predictor. Estimates in both models are adjusted for treatment regimen, age, sex, and HIV status. The reference group for both logistic models is disease class 0.

### 4.2 Disease Severity in the TRUST Study

The TRUST study was a prospective observational cohort of individuals initiating treatment for drug-susceptible TB at a clinic in Worcester, Western Cape Province, South Africa [17]. We include data on 351 participants enrolled between 2017 and 2022. Using clinical and laboratory measures collected at enrollment (Table 3) and their corresponding clinical states, we applied TB-STATIS to predict a disease severity class for each participant. We correlated predicted severity with 8-week culture status, a proxy for TB treatment response, and examined the relative frequencies of attributes known to associate with advanced TB across severity classes. Further details and additional results are provided in the Supplementary Material.

**Table 3.** Clinical measures in the TRUST cohort and estimated disease sequence from TB-STATIS.

| Data source | Clinical Measure | Clinical State | $P(x_i\vert x_i^*)$ | $\hat{S}$ |
| --- | --- | --- | --- | --- |
| Chest x-ray | Cavitary disease | Yes | 0.95 | 3 |
|  |  | No |  | 0 |
|  | Infiltrates | Bilateral | 0.95 | 4 |
|  |  | Unilateral |  | 1 |
|  |  | None |  | 0 |
| Sputum | Smear microscopy | +++ | 0.95 | 7 |
| | | Positive ≤ ++ | | 2 |
|  |  | Negative |  | 0 |
| Symptoms | Cough | Yes | 0.85 | 1 |
|  |  | No |  | 0 |
|  | Fever | Yes | 0.85 | 6 |
|  |  | No |  | 0 |
|  | Unexplained weight loss | Yes | 0.85 | 1 |
|  |  | No |  | 0 |
|  | Night sweats | Yes | 0.85 | 5 |
|  |  | No |  | 0 |

We estimated 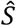 to have seven disease classes using our observed data and the specified values for 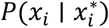 (Table 3). Using 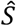, we predicted a disease class for each participant. The distribution of disease classes is left-skewed, with earlier stages occurring least frequently (Figure 6A). We observe a positive trend in log odds ratios between disease class and 8-week culture positivity (Figure 6B). Additionally, higher disease classes are associated with lower likelihood of HIV infection, higher BMI, and greater likelihood of drug use, factors known to correlate with advanced TB at presentation (Figure S10).

**Figure 6.**
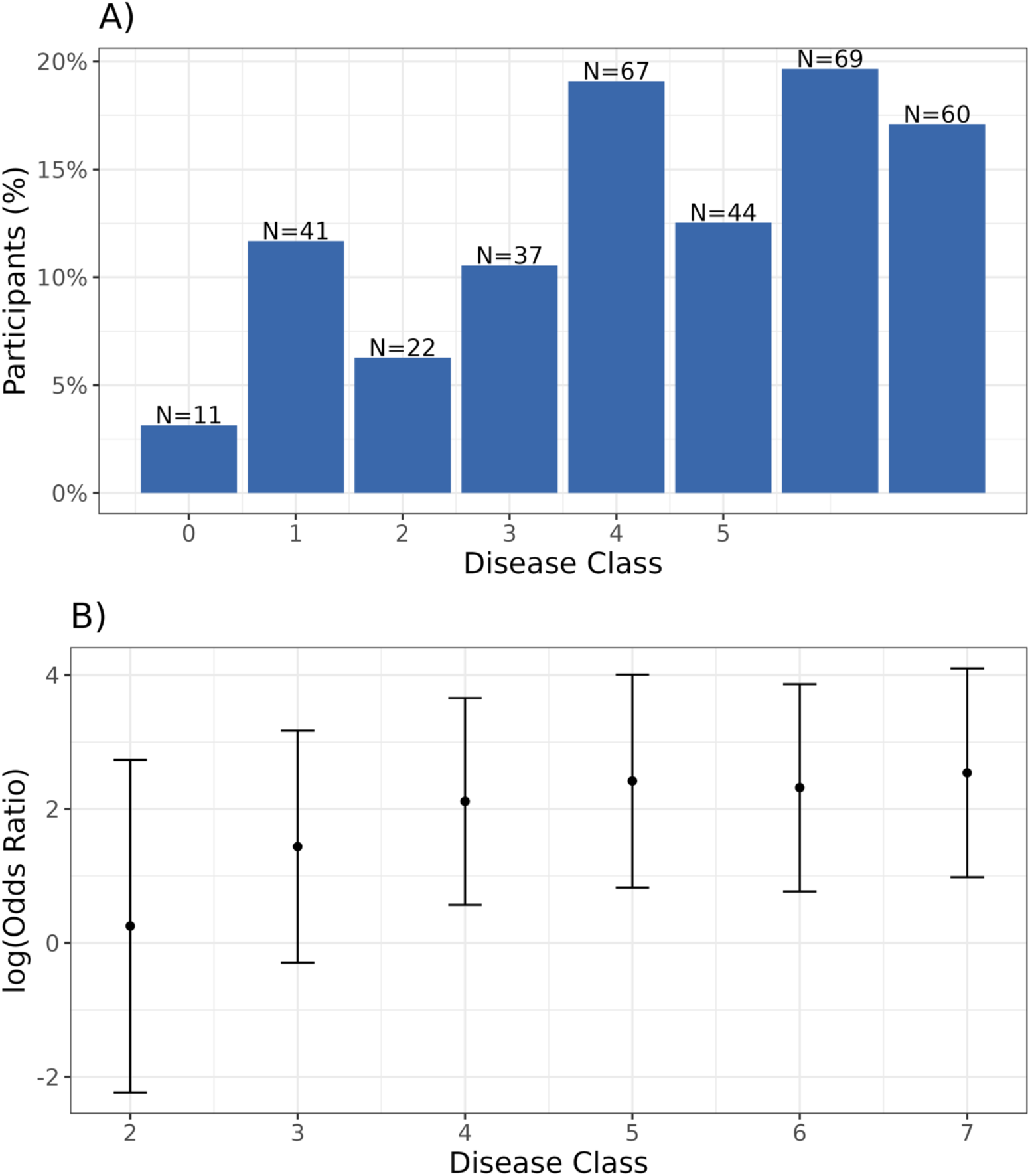
Disease severity classes generated by TB-STATIS and association with TB sterilization in the TRUST cohort. Panel A shows the distribution of predicted disease severity classes. Panel B shows the log odds ratios and 95% confidence intervals for the logistic regression model with 8-week culture positivity as the outcome and disease severity as a categorical predictor. Estimates are adjusted for age, sex, and HIV status. The reference group for the logistic model is the collapsed group of disease classes 0 and 1.

## 5 Concluding Remarks and Further Research

We developed a novel data-driven approach for classifying TB disease severity to better capture the wide spectrum of disease phenotypes observed at diagnosis. We introduce TB-STATIS as a flexible, statistically grounded method that classifies TB disease severity using only cross-sectional data and without requiring knowledge of clinical outcomes during estimation. In simulation, we demonstrate that our model accurately recovers an ordered sequence of disease-severity classes. Our approach was motivated by two extensions of the EBM: the model by Parker et al., which allows multiple events to occur simultaneously within a disease stage [11], and the model by Young et al., which accommodates ordinal data [12]. We integrate both frameworks and remove the need for control data, a requirement of Young et al. This addresses two key needs for TB data: handling ordinal clinical measures such as smear grade, Xpert Ultra cycle-threshold categories, and chest X-ray severity scores, and accommodating the reality that control data from individuals without TB are often unavailable.

We applied our model to TB data from a clinical trial and an observational cohort. Estimated disease severity classes correlated with TB sterilization in both the REMox trial and the TRUST study. In the TRUST study, severity class was also associated with HIV, BMI, and smoked drug use, factors previously linked to advanced TB [18]. These results suggest that our data-driven approach produces clinically meaningful severity classifications. Correlations with time to sterilization were weak and did not align with treatment outcomes, likely because the dataset primarily included individuals with very advanced disease. Future work should evaluate TB-STATIS using data with a broader distribution of disease severity, such as from low-burden settings or active case-finding programs.

Existing methods for classifying TB disease severity often do not utilize all diagnostic data collected at diagnosis. In clinical trials, stratification typically relies on cavitary disease on chest X-ray or sputum-based criteria like smear positivity [3,5,19,20]. Some severity scores, such as the Karnofsky Performance Score and Bandim TBscore, incorporate symptoms and correlate with treatment outcomes but exclude chest imaging or bacillary burden [21,22]. Melendez et al. developed a machine-learning framework integrating computer-aided chest X-ray scoring with clinical data (HIV status, mid-upper arm circumference, lung auscultation) but omitted bacillary burden and symptoms [23]. Imperial et al. proposed a risk stratification algorithm using HIV status, sex, smear grade, cavitary disease, BMI, and month 2 culture status, which limits stratification at diagnosis [24]. Ghanem et al. examined radiological and clinical measures, identifying an optimal model including percent lung involvement, age, sex, and smear grade for predicting unfavorable outcomes [25].

TB-STATIS captures disease severity across multiple dimensions. While we illustrate the method using symptoms, sputum bacillary burden, and chest X-ray, additional data such as detailed imaging outputs, lung function measures, inflammation markers, and rapid or non-sputum diagnostics can also be incorporated. A key strength of our approach is that it does not require a fixed set of variables or training on clinical outcomes, which is advantageous given global variability in diagnostic algorithms and research versus surveillance data. TB-STATIS also leverages prior knowledge of the TB disease spectrum through its EBM-based framework, unlike standard clustering or classification methods. Simulations show that it can effectively classify disease severity using up to 12 clinical states, providing a more refined understanding of TB severity.

Our method has limitations. TB disease progression is not linear, and severity measures can fluctuate. Some individuals may even “self-cure” and never be diagnosed. TB-STATIS focuses solely on classifying disease severity at diagnosis to stratify individuals by risk of poor outcomes; it does not infer disease trajectories or hypothetical progression without treatment. We assume the observed data are sufficient to classify severity, and if a person’s severity decreases by diagnosis, being placed in a lower class is appropriate. We also assume all individuals follow the same sequence of clinical states as severity increases. While this may hold at the population level, subgroups—such as previously treated patients, those with drug-resistant TB, or individuals with HIV—may follow different sequences. In such cases, we recommend fitting TB-STATIS separately for each subgroup. Future work could estimate disease subtypes directly from the data, as done with other EBM iterations [12,26].

TB-STATIS assumes all clinical measures are independent, which may not hold for TB, as diagnostics are often correlated (e.g., smear positivity and chest X-ray severity). In simulations with correlated measures, TB-STATIS still performed well with up to eight clinical states when accuracy was at least 0.85. Another limitation is the need to specify the accuracy of each measure; although validated diagnostics are expected to be highly accurate and near 1, true values are unknown and may vary, particularly for self-reported symptoms. Sensitivity analyses show the model remains robust even when accuracy is misspecified by 0.1. Future work incorporating uncertainty in measure accuracy and correlations between inputs could provide a more realistic, flexible model of the data generation process.

No standard method exists to assess TB disease severity, yet stratification at diagnosis and treatment initiation is essential. In clinical trials for shorter regimens, severity is increasingly used to tailor treatment, while in observational studies it often serves as a confounder when analyzing outcomes. TB programs also rely on severity assessment to identify high-risk patients and guide monitoring. We present a data-driven approach integrating multiple diagnostic measures at TB diagnosis to classify disease severity, demonstrating its clinical utility by correlating predicted classes with outcomes in two TB cohorts.

## 6 Software

The methods described here are implemented in the R package ‘tbSTATIS’ (https://github.com/samalatesta/tbSTATIS). The code used to generate all results in this article is also publicly (https://github.com/samalatesta/tbSTATISpaper).

## Data availability

The methods we developed and present in this article are implemented in the R package tbSTATIS which is available on GitHub at https://github.com/samalatesta/tbSTATIS. The code used to produce all results reported in this article is also available on GitHub at https://github.com/samalatesta/tbSTATISpaper. Data from the REMox trial are publicly available and were obtained from Platform for Aggregations of Clinical TB Studies. Data from the TRUST study used in the preparation of this article are available upon request.

## Acknowledgements

Data from the REMox trial used in the preparation of this article were obtained from Platform for Aggregations of Clinical TB Studies. The Platform for Aggregations of Clinical TB Studies initiative is a public private partnership launched in May 2015 by Critical Path Institute (C-Path), the Special Programme-for Research and Training in Tropical Diseases (TDR), the Global Alliance for TB Drug Development (TB Alliance), and St. George’s, University of London.

## References

1. Dartois VA, Rubin EJ. Anti-tuberculosis treatment strategies and drug development: challenges and priorities. Nat Rev Microbiol. 2022;20(11):685–701. doi:10.1038/s41579-022-00731-y

2. Motta I, Boeree M, Chesov D, et al. Recent advances in the treatment of tuberculosis. Clinical Microbiology and Infection. 2024;30(9):1107–1114. doi:10.1016/j.cmi.2023.07.013

3. Dorman SE, Nahid P, Kurbatova EV, et al. Four-Month Rifapentine Regimens with or without Moxifloxacin for Tuberculosis. New England Journal of Medicine. 2021;384(18):1705–1718. doi:10.1056/NEJMoa2033400

4. Chang VK, Imperial MZ, Phillips PPJ, et al. Risk-stratified treatment for drug-susceptible pulmonary tuberculosis. Nat Commun. 2024;15(1):9400. doi:10.1038/s41467-024-53273-7

5. Imperial MZ, Nahid P, Phillips PPJ, et al. A patient-level pooled analysis of treatment-shortening regimens for drug-susceptible pulmonary tuberculosis. Nat Med. 2018;24(11):1708–1715. doi:10.1038/s41591-018-0224-2

6. Esmail H, Macpherson L, Coussens AK, Houben RMGJ. Mind the gap – Managing tuberculosis across the disease spectrum. eBioMedicine. 2022;78. doi:10.1016/j.ebiom.2022.103928

7. Oxtoby NP. Data-Driven Disease Progression Modeling. In: Colliot O, ed. Machine Learning for Brain Disorders. Neuromethods. Springer US; 2023:511–532. doi:10.1007/978-1-0716-3195-9_17

8. Fonteijn HM, Modat M, Clarkson MJ, et al. An event-based model for disease progression and its application in familial Alzheimer’s disease and Huntington’s disease. NeuroImage. 2012;60(3):1880–1889. doi:10.1016/j.neuroimage.2012.01.062

9. Young AL, Oxtoby NP, Daga P, et al. A data-driven model of biomarker changes in sporadic Alzheimer’s disease. Brain. 2014;137(9):2564–2577. doi:10.1093/brain/awu176

10. Venkatraghavan V, Bron EE, Niessen WJ, Klein S. Disease progression timeline estimation for Alzheimer’s disease using discriminative event based modeling. NeuroImage. 2019;186:518–532. doi:10.1016/j.neuroimage.2018.11.024

11. Parker CS, Oxtoby NP, Young AL, Initiative ADN, Alexander DC, Zhang H. Parsimonious EBM: generalising the event-based model of disease progression for simultaneous events. Published online March 11, 2024:2022.07.10.499471. doi:10.1101/2022.07.10.499471

12. Young AL, Vogel JW, Aksman LM, et al. Ordinal SuStaIn: Subtype and Stage Inference for Clinical Scores, Visual Ratings, and Other Ordinal Data. Front Artif Intell. 2021;4. doi:10.3389/frai.2021.613261

13. Lourenco HR, Martin OC, Stutzle T. Iterated Local Search. Published online February 23, 2001. Accessed May 16, 2024. http://arxiv.org/abs/math/0102188

14. Hamming RW. Error detecting and error correcting codes. The Bell System Technical Journal. 1950;29(2):147–160. doi:10.1002/j.1538-7305.1950.tb00463.x

15. Fagin R, Kumar R, Mahdian M, Sivakumar D, Vee E. Comparing Partial Rankings. SIAM J Discrete Math. 2006;20(3):628–648. doi:10.1137/05063088X

16. Gillespie SH, Crook AM, McHugh TD, et al. Four-Month Moxifloxacin-Based Regimens for Drug-Sensitive Tuberculosis. N Engl J Med. 2014;371(17):1577–1587. doi:10.1056/NEJMoa1407426

17. Myers B, Bouton TC, Ragan EJ, et al. Impact of alcohol consumption on tuberculosis treatment outcomes: a prospective longitudinal cohort study protocol. BMC Infect Dis. 2018;18(1):488. doi:10.1186/s12879-018-3396-y

18. Myers B, Carney T, Rooney J, et al. Smoked drug use in patients with TB is associated with higher bacterial burden. int j tuberc lung dis. 2023;27(6):444–450. doi:10.5588/ijtld.22.0650

19. Franke MF, Khan P, Hewison C, et al. Culture Conversion in Patients Treated with Bedaquiline and/or Delamanid. A Prospective Multicountry Study. Am J Respir Crit Care Med. 2021;203(1):111–119. doi:10.1164/rccm.202001-0135OC

20. Paton NI, Cousins C, Suresh C, et al. Treatment Strategy for Rifampin-Susceptible Tuberculosis. New England Journal of Medicine. 2023;388(10):873–887. doi:10.1056/NEJMoa2212537

21. Schag CC, Heinrich RL, Ganz PA. Karnofsky performance status revisited: reliability, validity, and guidelines. J Clin Oncol. 1984;2(3):187–193. doi:10.1200/JCO.1984.2.3.187

22. Wejse C, Gustafson P, Nielsen J, et al. TBscore: Signs and symptoms from tuberculosis patients in a low-resource setting have predictive value and may be used to assess clinical course. Scandinavian Journal of Infectious Diseases. 2008;40(2):111–120. doi:10.1080/00365540701558698

23. Melendez J, Sánchez CI, Philipsen RHHM, et al. An automated tuberculosis screening strategy combining X-ray-based computer-aided detection and clinical information. Sci Rep. 2016;6(1):25265. doi:10.1038/srep25265

24. Imperial MZ, Phillips PPJ, Nahid P, Savic RM. Precision-Enhancing Risk Stratification Tools for Selecting Optimal Treatment Durations in Tuberculosis Clinical Trials. Am J Respir Crit Care Med. 2021;204(9):1086–1096. doi:10.1164/rccm.202101-0117OC

25. Ghanem M, Srivastava R, Ektefaie Y, et al. Percent of lung involved in disease on chest X-ray predicts unfavorable treatment outcome in pulmonary tuberculosis. Published online August 20, 2024:2024.08.19.24311411. doi:10.1101/2024.08.19.24311411

26. Young AL, Marinescu RV, Oxtoby NP, et al. Uncovering the heterogeneity and temporal complexity of neurodegenerative diseases with Subtype and Stage Inference. Nat Commun. 2018;9(1):4273. doi:10.1038/s41467-018-05892-0

